# Accuracy and Generalizability of an Open-Source Deep Learning Model For Facial Bone Segmentation on CT and CBCT Scans

**DOI:** 10.64898/2025.12.28.25343101

**Authors:** Nikolaos Gkantidis, Mohammed Ghamri, Gauthier Dot

## Abstract

**Aim:** To evaluate the accuracy and generalizability of DentalSegmentator, an open-source deep learning tool, for automated segmentation of skeletal facial surfaces from computed tomography (CT) scans acquired under different imaging conditions.

**Materials and Methods:** Ten human skulls were scanned using a CT scanner and three cone beam CT (CBCT) protocols (including an ultra-low-dose protocol) on two CBCT devices. High-accuracy reference surface models were acquired using an optical scanner. CBCT an CT scans were segmented automatically using DentalSegmentator. Three facial regions (forehead, zygomatic process, maxillary process) were defined on each model for quantitative assessment. Accuracy was measured as the mean absolute distance (MAD) and the standard deviation of absolute distances (SDAD) between segmented and reference models after best-fit superimposition.

**Results:** Repeated segmentations were identical, confirming perfect reproducibility. Across all acquisition settings and regions, DentalSegmentator produced highly accurate skeletal surface models, with an overall MAD of 0.088 mm (IQR 0.073) and SDAD of 0.061 mm (IQR 0.028). Significant but small differences were detected between imaging systems (MAD: p < 0.001; SDAD: p = 0.003), with CT scans showing slightly reduced trueness compared with CBCT images.

**Conclusion:** The open-source DentalSegmentator tool produced accurate skeletal facial surface segmentations across diverse CT and CBCT settings, demonstrating excellent generalizability, including under low-radiation conditions. Minor differences in trueness between imaging systems were small and unlikely to impact clinical or research use.

**Clinical Significance:** Deep learning offers a robust foundation for automated 3D craniofacial surface extraction, supporting broader adoption of AI-driven workflows in both clinical and research contexts.

## Introduction

Digital dentistry and craniofacial surgery workflows now depend greatly on digital 3D models created from multi-slice computed tomography (CT) or cone-beam computed tomography (CBCT) scans. Models depicting skeletal facial surfaces are particularly important for computer-assisted surgical planning [1], virtual patient generation [2], or the superimposition of craniofacial structures over time [3]. After scan acquisition, a critical step in the workflow is the accurate delineation of anatomical structures through segmentation. Over the past five years, segmentation methods for dento-maxillo-facial (DMF) CT and CBCT scans have shifted substantially. Although manual segmentation remains the reference standard, artificial intelligence (AI), particularly deep learning–based approaches, has shown strong potential for automating this process [4–7]. Accordingly, clinicians now have access to open-source and commercial automated solutions for segmenting all anatomical components of the DMF complex [2,8–10].

Given the surge in AI-based segmentation solutions, it is crucial to evaluate them with methodological rigor and adhere to guidelines such as DENTAL-AI for planning, conducting, and reporting studies [11]. A recent review of AI-based DMF segmentation concluded that although research papers report very encouraging results that are on par with or even better than those of expert-based segmentation, methodological flaws in the included studies may affect the results [5]. High-quality studies addressing real clinical problems or advances in AI methodology are needed [12].

The accuracy of AI-based segmentation reflects the trueness and precision of the resulting 3D models [13] and is typically assessed by comparison with expert-based reference segmentations. This approach is required because obtaining a hard ground truth, such as the true skeletal facial surface, is not feasible in living subjects due to ethical and safety constraints, including excessive radiation exposure or invasive procedures. However, reference annotations rely on manual segmentations or manual correction of automated outputs and are therefore subject to operator variability [11]. To our knowledge, no previous study has evaluated AI-based DMF segmentation using a hard ground truth. An *ex vivo* design using dry cadaveric specimens is therefore the only feasible approach for accurately representing true anatomy [14]. Current best practice involves scanning hydrated specimens with a high-accuracy optical surface scanner [15] and embedding them in soft tissue simulants during tomographic acquisition to approximate clinical conditions [16].

The generalizability of an AI-based segmentation model refers to its ability to maintain accuracy on new data, including data acquired with previously unseen equipment, acquisition protocols, or image resolutions [17]. Ideally, it is evaluated using an external test set comprising independent data from other institutions [18]. Although the use of external test sets has increased in recent studies, few investigations assessed how equipment and acquisition parameters affect automated segmentation performance [19]. Moreover, the effect of low-radiation protocols on segmentation accuracy remains unclear [20].

The clinical acceptability of AI-based segmentation depends on the use of clinically relevant and meaningful performance metrics [12]. Most DMF segmentation studies primarily report the Dice coefficient [1,5]. While useful for model comparison, this metric has limited clinical interpretability and may not reflect clinical relevance [21]. Though normalized surface distance has been proposed as a clinically relevant alternative, it is still derived from engineering and may lack clinical significance [22]. Other methods are often used in clinical studies. For example, distance metrics between 3D surface models or colour-coded distance maps are commonly used to evaluate 3D superimposition results [23,24]. The use of these methods in evaluating AI-based segmentation has yet to be investigated. A methodological study applying such clinically meaningful methodologies will strengthen the foundation of deep learning-based methods [12].

Therefore, this *ex vivo* study aimed to evaluate the accuracy and generalizability of DentalSegmentator, an open-source AI-based segmentation method [8], for reconstructing 3D facial skeletal surface models. Accuracy was assessed by comparison with direct surface scans used as ground truth. Generalizability was evaluated across multiple CT and CBCT scanners and acquisition protocols, including an ultra-low-dose protocol. Clinically relevant measures, including distance maps, were applied under the assumption of no significant differences between segmentation results and the ground truth across scanners and protocols.

## Materials and Methods

### Ethical approval and informed consent

Ethical approval for this study was obtained from the Research Ethics Committee of the Dental School, National and Kapodistrian University of Athens (Protocol No. 335; initial approval 02/05/2017, renewed 16/11/2021). All procedures adhered to applicable guidelines and regulatory standards. Because the specimens were anonymized and their donors could not be identified, the committee issued a waiver for informed consent. Handling of all human tissues complied with the corresponding local legal requirements.

### Sample

This study utilized ten human dry skulls obtained from the Municipal Cemetery of Serres, Greece, with formal approval from the local authorities (Municipality of Serres, Protocol No. 4044/12.07.2018). The specimens belonged to unidentified individuals who had died 8 to 12 years prior to sample collection. At the time of retrieval, no claims had been made by relatives, and the skulls had been buried collectively according to cemetery protocol. Each skull was visually inspected by two evaluators to confirm that it represented a fully developed adult cranium with no major defects, pathological conditions, or significant age related deterioration.

### Data acquisition

#### Optical reference models

All ten specimens were scanned using a structured-light 3D surface scanner (Artec Space Spider, Artec3D, Luxembourg; Software: Artec Studio 12, Version 12.1.6.16) to acquire high-resolution reference surface models that comprised the ground truth models for the study. To ensure dimensional stability between optical and tomographic scanning, all skulls were hydrated by immersion in tap water for 15 minutes before scanning. Scanning was performed immediately after removal from water, following gentle patting with tissue paper. The water temperature ranged between 22–25 °C, with an approximate pH of 7.5. The optical scanner precision [25], as well as the effect of hydration on the anatomical form of facial skeletal bones [16] has been thoroughly evaluated previously on the same specimens and the subsequent surface models have been used for the present study. Raw optical scan data were processed in Artec Studio 16 (Version 16.0.5.114). Partial scans were merged into complete skull models through automated alignment or a combination of initial manual landmark approximation followed by automated registration. The resulting ten reference models were imported into Viewbox 4 software (dHAL Software, Kifisia, Greece) for subsequent comparisons with the tomographic surface based models.

#### Tomographic Data

The tomographic scans included in this study were performed on the same ten skulls. These were previously used in publications evaluating their trueness [14,26]. Soft tissue simulation was achieved using a 3D-printed head shell filled with water (PETG, MasterFill Premium PETG Pro, 3DHUB, Greece), aiming to similar hydration conditions to those during optical scanning. As previously published [26], each skull was scanned using four tomographic protocols, generating ten volumes per acquisition setting (total: 40 volumes):

1. CT scanner (Revolution CT 256, GE Healthcare, USA; 251 Hellenic Airforce Hospital, Athens, Greece). Tube voltage: 120 kV, tube current: 490 mA in the area of interest (automatically configured based on tissue mass and density), exposure time: 1 s, slice thickness: 0.625 mm, voxel size: 0.49 to 0.62 × 0.49 to 0.62 × 0.31 (interslice) mm, FOV: full head (displayed FOV: 25 cm).
2. CBCT scanner I (Newtom VGiMK4, Verona, Italy; Dental School, National and Kapodistrian University of Athens Greece). Tube voltage: 110 kV, tube current: 4–5 mA (automatically configured based on tissue mass and density), exposure time: 4 s, voxel size: 0.3 × 0.3 × 0.3 mm, FOV: ⌀15 × 15 cm.
3. CBCT scanner II—regular dose settings (Planmeca F, Planmeca Promax 3D Mid 2018, Helsinki, Finland; Digital Iatriki Apeikonisi, Athens, Greece). Tube voltage: 100 kV, tube current: 8 mA, exposure time: 12 s, voxel size: 0.2 × 0.2 × 0.2 mm, FOV: ⌀20 × 17 cm.
4. CBCT scanner II—ultra-low dose settings (Planmeca U). Tube voltage: 100 kV, tube current: 8 mA, exposure time: 6s, voxel size: 0.2 × 0.2 × 0.2 mm, FOV: ⌀20 × 17 cm.

All tomographic DICOM datasets were converted in Neuroimaging Informatics Technology Initiative (NIfTI) files using ITK-Snap software (v. 4.0.1, http://www.itksnap.org/) [27]. Those NIfTI files were automatically segmented by one operator (GD) using the publicly available nnU-Net DentalSegmentator weights with no additional pre- or post-processing [8,28]. The resulting segmentation masks were then imported as NifTI files into Viewbox 4 and automatically reconstructed as 3D surface models (STL) using the software’s built-in isosurface reconstruction (marching-cubes) algorithm.

### Reproducibility assessment

To evaluate reproducibility, all four tomographic images from two randomly selected skulls were segmented twice using the DentalSegmentator software. Given the fully automated nature of the segmentation process, identical results were expected across repeated runs. In the event of discrepancies, a detailed reproducibility analysis would be conducted. Colour-coded distance maps between best-fit approximated, repeatedly segmented surface models were generated to assess this outcome.

### Surface Model Superimposition and Trueness Assessment

A best-fit registration procedure was performed to superimpose each CBCT/CT derived 3D model onto its corresponding optical reference model. A variant of the iterative closest point (ICP) algorithm [29] was used with the following parameters: 100% estimated overlap of meshes, matching point to plane, exact nearest neighbour search, 100% point sampling, 50 iterations. The reference areas used for superimposition and the predefined measurement regions are illustrated in Figure 1.

**Figure 1.**
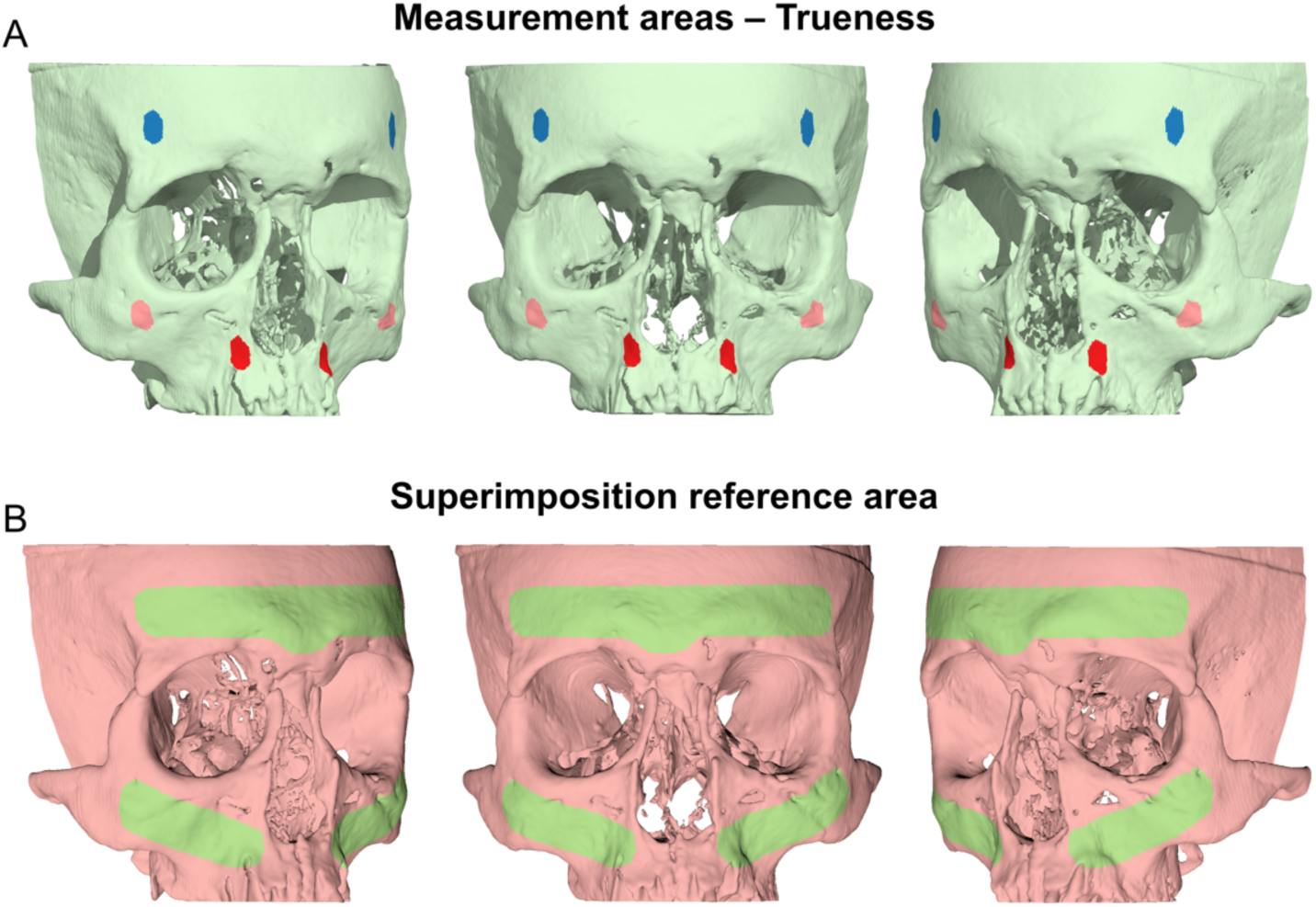
A. Three measurement areas defined in each skull through bilaterally selected mesh surfaces at the forehead (blue), the zygomatic process (pink), and the maxillary process (red) for trueness assessment, respectively. B. Reference area used for all surface-based superimpositions performed in the study (green).

Measurement areas were consistently placed on the ground truth models and included bilateral areas on the forehead, zygomatic processes, and maxillary complex (Figure 1). Each single selection covered an area of 60 mm^2^ and consisted of approximately 900 vertices. Bilateral areas were treated as single measurement units.

Trueness of the DentalSegmentator derived surface models was quantified by calculating the congruence between the superimposed models using Mean Absolute Distance (MAD) and Standard Deviation of Absolute Distances (SDAD). Both metrics were calculated for each of the three predefined measurement regions. Colour-coded distance maps were generated to visually illustrate deviations over the entire surface model. Perfect trueness corresponded to regions showing zero deviations.

### Statistical analysis

The IBM SPSS statistics for Windows (Version 31.0.IBM Corp., Armonk, NY, USA) was used for descriptive and comparative statistical analyses and associated graph generation. The significance level was set at 0.05. Data normality was assessed using the Shapiro–Wilk test and visual inspection of normality plots, revealing significant deviations for some variables. Therefore, non-parametric statistical methods were used.

Differences in the trueness of segmented facial surface models across the four acquisition settings were assessed using the Kruskal–Wallis test. If significant, pairwise comparisons were conducted with the Dunn’s test, applying Bonferroni correction. Analyses were performed both for the combined measurement areas and for each area individually.

## Results

### Reproducibility

All repeatedly segmented facial surface models from the tomographic scans (n=8: 4 acquisition settings x 2 skulls) through DentalSegmentator were identical to the original segmentations (Figure 2). Therefore, no further reproducibility assessment was performed.

**Figure 2.**
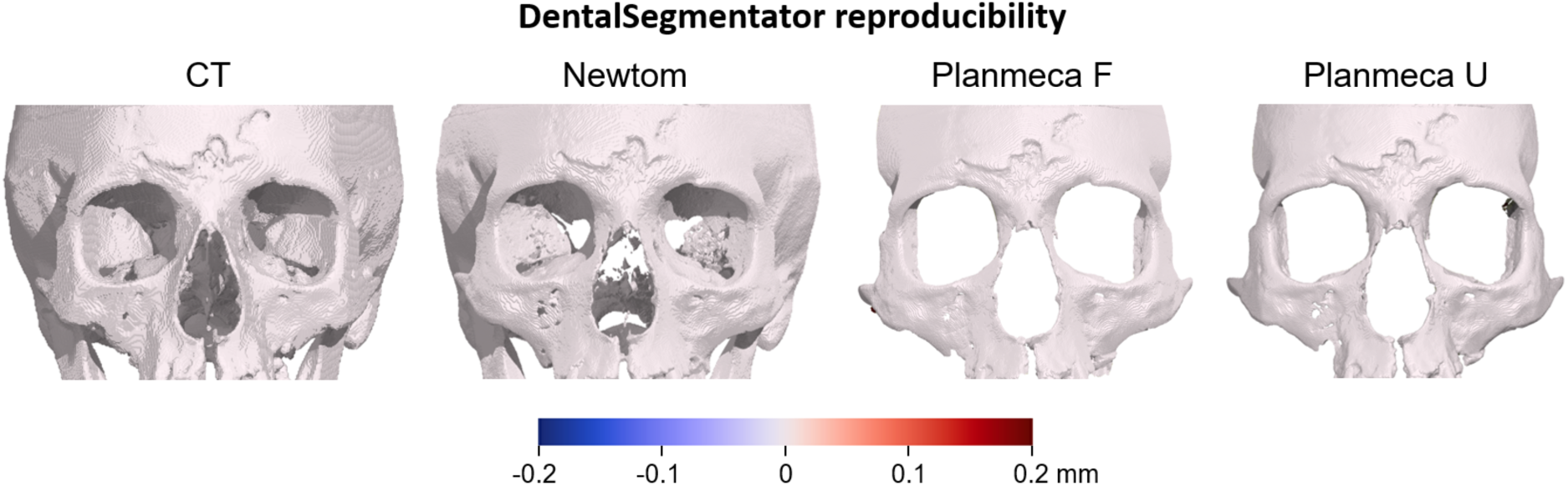
Colour-coded distance maps comparing repeatedly segmented surface models of a single skull generated using DentalSegmentator. The models retain their original spatial relationship within the source tomographic volume (prior to superimposition). The first two panels show part of the entire scanned skull, while the last two show only the region of interest assessed in this study.

### Trueness

Considering all acquisition settings and measurement areas, the overall trueness (MAD) of DentalSegmentator derived skeletal facial surfaces was 0.088 mm (IQR: 0.073), with an SDAD of 0.061 mm (IQR: 0.028). There were significant differences between acquisition settings in MAD values depicting trueness (Kruskal-Wallis Test: p<0.001), as well as in the respective SDAD values (Kruskal-Wallis Test: p=0.003), although these were of limited magnitude (Figure 3). The segmentation of CT scans provided slightly less accurate outcomes than those of the Planmeca scans. Detailed descriptive statistics and pairwise comparison test are provided in Table 1.

**Figure 3.**
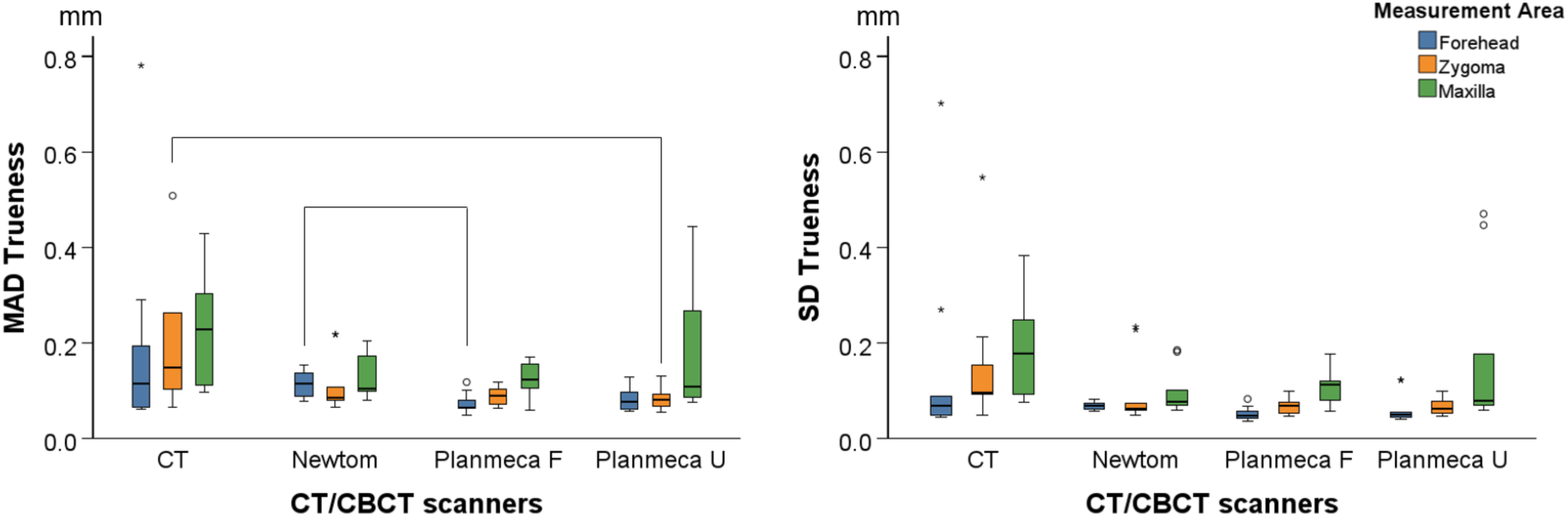
Box plots showing the trueness of the segmented facial surface models indicated by the distances of the tomographically derived models from the direct optical scans, following their best-fit approximation. The Mean Absolute Distances (MAD) and the standard deviations of the absolute distances (SD) between the superimposed surface models are shown. The lines connect variables that show significant differences (p<0.05) detected through Kruskal-Wallis, followed by Dunn’s test (Bonferroni adjusted). Outliers are shown as black circles (further from the median more than 1.5 times the IQR) and extreme outliers as asterisks.

**Table 1.** Trueness of the DentalSegmentator facial skeletal surface models indicated by MAD and SDAD values per tomographic acquisition setting and associated pairwise comparisons.

|  | MAD |  | SDAD |  |
| --- | --- | --- | --- | --- |
|  | *Median* | *IQR* | *Median* | *IQR* |
| **CT** | 0.148 | 0.169 | 0.095 | 0.140 |
| **Newtom** | 0.101 | 0.065 | 0.070 | 0.019 |
| **Planmeca F** | 0.090 | 0.053 | 0.067 | 0.045 |
| **Planmeca U** | 0.087 | 0.046 | 0.067 | 0.036 |
|  | *Pairwise Comparisons* |  | *Pairwise Comparisons* |  |
|  | *Sig.* | *Adj. Sig.<sup>a</sup>* | *Sig.* | *Adj. Sig.<sup>a</sup>* |
| **Planmeca F vs. Planmeca U** | 0.778 | 1.000 | 0.973 | 1.000 |
| **Planmeca F vs. Newtom** | 0.060 | 0.359 | 0.367 | 1.000 |
| **Planmeca F vs. CT** | <0.001 | 0.001 | 0.001 | 0.008 |
| **Planmeca U vs. Newtom** | 0.110 | 0.658 | 0.385 | 1.000 |
| **Planmeca U vs. CT** | <0.001 | 0.003 | 0.002 | 0.009 |
| **Newtom vs. CT** | 0.062 | 0.375 | 0.021 | 0.127 |

Asymptotic significances (2-sided tests) are displayed. The significance level is 0.05.
a. Significance values have been adjusted by the Bonferroni correction for multiple tests.

When each measurement area was analysed as a single variable, there were only minor and very few statistically significant differences in trueness (MAD of the best-fit approximated models) between acquisition settings (Forehead, Newtom vs Planmeca F: p=0.026; Zygoma, CT vs Planmeca U: p=0.015). Similarly, for the SDAD there were no pairwise differences between acquisition settings although the Kruskal-Wallis test was significant for the Forehead area (p=0.024) (Figure 3).

The colour-coded distance maps between the best-fit approximated segmented and directly scanned facial surface models revealed consistent outcomes within and between acquisition settings, with deviations from the true model only locally reaching a maximum of 0.5 mm (Figure 4). Large deviations were often present at the sides of the skull, located distant to the used superimposition reference area.

**Figure 4.**
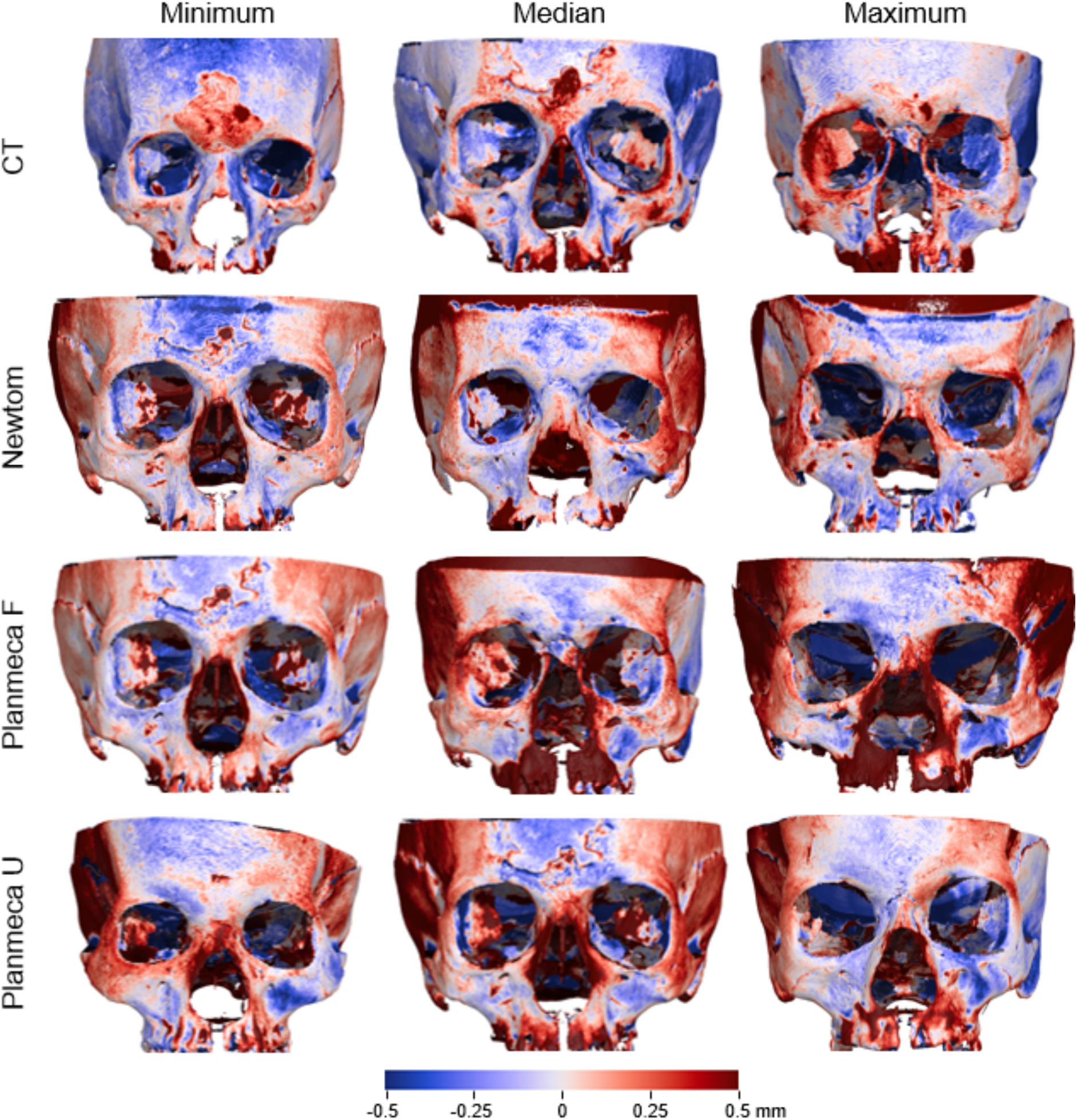
Colour-coded distance maps between best-fit approximated segmented surface models and directly obtained models through an optical surface scanner, representative of the minimum, average, and maximum deviations in trueness, detected in the sample for each acquisition setting.

## Discussion

This *ex vivo* study evaluated the accuracy and generalizability of DentalSegmentator, an open-source deep learning model for automatic segmentation of skeletal facial surfaces from CT and CBCT scans [8]. Overall, the tool produced highly accurate 3D facial bone models, with median mean absolute deviations (MAD) below 0.1 mm and standard deviations of absolute distances (SDAD) around 0.06 mm when compared with high-precision optical surface scans [25] used as ground truth. Although differences among tomographic scanners and acquisition settings, including the ultra-low-dose CBCT protocol, occasionally reached statistical significance, their magnitude was minimal and unlikely to be clinically meaningful. Colour-coded distance maps confirmed that local deviations rarely exceeded 0.5 mm and were mostly confined to lateral regions away from the superimposition reference area. These findings suggest that, for facial skeletal surfaces, DentalSegmentator is not a major source of geometric error and can be considered suitable for demanding clinical and research applications.

The present results are consistent with, and extend, previous work on the accuracy of tomographically derived facial surface models. In a closely related *ex vivo* study using the same skulls and a visually defined single-threshold segmentation approach, the overall trueness of CT- and CBCT-derived facial surfaces was approximately 0.12 mm, with local deviations again reaching up to 0.5 mm and no relevant differences between scanners or protocols [14,26]. In the current investigation, the overall MAD of the DentalSegmentator-derived surfaces (0.088 mm) was slightly lower than those previously reported for manual or semi-automatic threshold-based segmentations of the same tomographic data, while the spatial pattern and magnitude of local deviations remained similar. These findings suggest that, under realistic imaging conditions, geometric accuracy is primarily constrained by the tomographic acquisition, with deep learning–based segmentation performing at least as well as, and in this study slightly better than, manual or semi-automatic approaches.

These observations align well with the original multicentre evaluation of DentalSegmentator on clinical CT and CBCT datasets, where the model achieved high Dice similarity coefficients and normalized surface distances for multiple DMF structures [8]. In that work, segmentation quality was judged primarily through voxel-level overlap metrics against expert segmentations. By contrast, the present *ex vivo* study uses an independent physical ground truth and clinically interpretable surface distances, thereby complementing and strengthening the evidence base for the model. Together, these data support the notion that DentalSegmentator provides robust and geometrically accurate segmentations across a variety of scanners and protocols, at least for the facial skeleton.

From a generalizability perspective, the inclusion of one CT and two CBCT units, as well as a reduced-dose CBCT acquisition, is noteworthy. External benchmarks in medical image segmentation, such as the Medical Segmentation Decathlon and more recent large-scale frameworks, have demonstrated that robust models must cope with multicentre, multi-device, and protocol heterogeneity to perform reliably in real-world settings [30]. The impact of scanner type and acquisition parameters on the results of DMF AI segmentation has recently been quantified in a large-scale study demonstrating that, for a given model, segmentation accuracy can vary by up to 25% in Dice score depending on the scans characteristics [19]. The present findings indicate that DentalSegmentator maintains sub-0.15 mm median MAD across all tested settings, with the lowest accuracy associated with the CT scanner and slightly better outcomes for the CBCT devices, including the ultra-low-dose configuration. This pattern mirrors earlier *ex vivo* work showing that ultra-low-dose CBCT protocols can preserve the trueness of facial surface models at levels comparable to regular-dose protocols and CT, thereby supporting further dose reduction in clinical practice [14,26].

The use of an *ex vivo* experimental design on dry skulls embedded in soft-tissue simulants is an important methodological strength [16]. This approach is currently regarded as the most rigorous way to obtain a ground truth for skeletal surface geometry in radiology research, because it decouples segmentation error from imaging limitations and allows direct comparison with high-precision surface scans [16,25]. In the present study, ground-truth models were acquired with a structured-light scanner whose precision has been shown to be on the order of tens of micrometres and whose model-generation error is negligible [25]. Hydration of the skulls and embedding in water-filled head shells followed previously validated protocols that mimic soft-tissue attenuation while inducing only limited geometric changes in the skeletal anatomy [16]. This combination of high-fidelity reference data and clinically realistic imaging conditions provides a solid basis for evaluating AI segmentation accuracy.

Another key contribution of this study lies in the choice of the evaluation metrics. Most AI-based segmentation studies in DMF imaging rely on volumetric overlap measures such as Dice or Jaccard coefficients, often complemented by global surface distances such as normalized surface distance, which are useful for benchmarking algorithms but can be difficult to interpret clinically [21,31]. Recent methodological work in medical imaging has emphasized that metric choice should reflect the clinical task and that overreliance on single, global metrics can obscure clinically relevant failure cases. In line with recommendations for AI in dental research, which call for transparent reporting and task-specific performance measures [11], the present study adopts local mean absolute distances and colour-coded surface maps as primary outcomes. These metrics are widely used to assess craniofacial superimposition and 3D treatment changes, and their magnitude can be directly related to clinically acceptable thresholds for diagnosis, treatment planning, or 3D printing [32,33].

From a clinical standpoint, the observed level of error appears acceptable for most applications involving facial skeletal surfaces. Previous accuracy studies on CT- and CBCT-derived craniofacial models have reported surface deviations in the range of 0.1–0.2 mm for well-chosen acquisition protocols, with local deviations occasionally reaching 0.5 mm in areas of thin bone or at the periphery of the field of view [14,26]. The present findings fall within or slightly below this range, despite the fully automatic nature of the segmentation process. Such accuracy is likely sufficient for virtual treatment planning, orthognathic surgery simulations, assessment of facial morphology, 3D printing CAD-CAM applications and longitudinal 3D superimposition, where sub-millimetre discrepancies are typically considered negligible relative to biological variability and growth- or treatment-related changes [34–36]. However, for high accuracy-sensitive procedures, such as the design of cutting guides or patient-specific implants in regions of very thin cortical bone, careful visual inspection of the segmentation remains advisable.

The present study also complements recent work investigating the performance of DentalSegmentator and other deep learning tools for tooth segmentation in challenging clinical scenarios. In a study focusing on patients with multiple impacted teeth, DentalSegmentator and a commercial cloud-based solution achieved high normalized surface distances and low failure rates, outperforming two other commercial tools in several metrics [9]. Together with the current results on skeletal surfaces, these findings support the broader use of open-source deep learning models as reliable building blocks for constructing full virtual patients and for integrating AI into advanced orthodontic and orthognathic workflows.

Despite its strengths, this study also has limitations. First, the sample size was limited to ten adult skulls, which restricts the variability of craniofacial morphology, bone density, and potential artefacts. While this is acceptable for a methodological validation study, larger and more heterogeneous datasets, ideally including pathological cases and different age groups, are needed to further probe model robustness. Second, the *ex vivo* design, although optimal for ground-truth acquisition, does not capture soft-tissue motion or patient positioning variability that are common in clinical imaging [37]. Third, because only one deep learning model and restricted set tomographic scanners and acquisition protocols were assessed, the results may not be fully generalizable to all available systems or settings. Finally, this study thoroughly evaluated facial skeletal surface segmentation results but did not evaluate other anatomical parts such as the anterior cranial base, the mandible or the teeth.

Future research should address these limitations by extending the evaluation to *in vivo* datasets with rigorously curated reference segmentations and by incorporating a broader range of scanners, voxel resolutions, and artefact conditions. In addition, comparative analyses involving diverse AI architectures, including medically adapted variants of foundation models or “segment-anything” frameworks [38], may help clarify their potential advantages for CBCT segmentation tasks. The integration of uncertainty estimation and automated quality-control procedures could further assist in identifying cases in which model outputs may be unreliable and therefore warrant manual verification. Finally, linking segmentation accuracy to downstream clinical tasks—for example, quantification of skeletal changes, prediction of surgical outcomes, or fabrication of guides and splints—would provide further assessment of clinical utility, consistent with recent calls to move beyond purely technical evaluations [18].

## Conclusions

This *ex vivo* investigation demonstrates that DentalSegmentator can accurately reconstruct skeletal facial surface models from CT and CBCT scans acquired under realistic clinical conditions, with errors that are small in magnitude and comparable to, or lower than, those introduced by the imaging process itself. By integrating a rigorous ground-truth approach, multi-scanner evaluation, and clinically meaningful evaluation metrics, this study provides robust evidence supporting the use of open-source deep learning–based segmentation tools in craniofacial imaging and digital dentistry.

## Data availability

All generated and processed measurement data are included within the main article and the Supplementary Dataset. Due to ethical and privacy restrictions related to the sensitive nature of the specimens, raw surface and tomographic data are not publicly available but may be provided by the corresponding author upon reasonable request.

## Author contributions

Conceptualization, N.G. and D.G.; Methodology, N.G., M.G. and D.G.; Software, N.G. and D.G.; Validation, N.G., M.G. and D.G.; Formal analysis, N.G.; Investigation, N.G., M.G. and D.G.; Resources, N.G., M.G. and D.G.; Data curation, N.G., M.G. and D.G.; Writing – original draft, N.G., M.G. and D.G.; Writing – review & editing, N.G., M.G. and D.G.; Visualization, N.G. and M.G.; Supervision, N.G. and D.G.; Project administration, N.G.; Funding acquisition, N.G. All authors have read and approved the final manuscript.

## Acknowledgements

This work was supported by the Swiss Dental Association through research grants (Protocol Nr. 335-21 and 345-23) awarded to author N. Gkantidis; and by the European Orthodontic Society, in the context of the W J B Houston Scholarship Award that was granted to author N. Gkantidis.

